# TRANSIENT ALTERATIONS IN THALAMO-CEREBELLAR FUNCTIONAL CONNECTIVITY IN PREMANIFEST HUNTINGTON’S DISEASE

**DOI:** 10.1101/2025.01.15.25320232

**Authors:** Melanie A. Morrison, Jingwen Yao, Radhika Bhalerao, Angela Jakary, Julia Glueck, Theresa Driscoll, Michael D. Geschwind, Alexandra B. Nelson, Katherine L. Possin, Christopher P. Hess, Janine M. Lupo

## Abstract

**Background:** There are no disease modifying therapies for Huntington’s disease (HD), a rare but fatal genetic neurodegenerative condition. To develop and test new management strategies, a better understanding of the mechanisms underlying HD progression is needed. Aberrant changes in thalamo-cortical and striato-cerebellar circuitry have been observed in asymptomatic HD, along with transient enlargement of the dentate nucleus.

**Purpose:** To evaluate the relationship between thalamo-cerebellar connectivity and HD progression.

**Study Type:** Prospective and retrospective.

**Population:** Patients with HD and healthy controls from a single-center dataset (n=34), and patients from the public TRACK-HD dataset (n=91).

**Field strength/Sequence:** 3T and 7T.

**Assessment:** Thalamo-cerebellar connectivity was compared across patients and controls and related to motor scores and predicted years to symptom onset. Cross-sectional findings were validated within-patient by mapping changes in individual connectivity over time. HD effects on cognitive performance were also explored and related to connectivity.

**Statistical Tests:** Kruskal-Wallis with post hoc Dunn’s tests and Pearson correlations (p_significant_<0.05).

**Results:** In the 7T cohort, significant premanifest and control group differences in thalamo-dentate connectivity were observed (p_Dunn_<0.05, η^2^=.19-.22), with manifest HD connectivity approaching normative values. Thalamic connectivity with the dentate nucleus and anterior cerebellum also correlated with years to onset (p_Den_=0.06, r=0.42, p_Ant_<0.05, r=-0.45), together indicating potential transient functional alterations in premanifest HD. Similar patterns were observed between connectivity (thalamus to dentate nucleus and anterior lobe) and cognitive performance scores across all subjects (p<0.05, r_Den_=-0.17, r_Ant_=-0.18). In the premanifest TRACK-HD cohort, connectivity of multiple thalamo-cerebellar connections correlated with years to onset, revealing distinct patterns for patients with low versus high motor scores, again indicative of potential transient alterations. Exploratory non-parametric regression of serial imaging data further supported these findings.

**Data conclusion:** Transient changes in thalamo-cerebellar connectivity are seen in premanifest HD with increasing progression. More studies are needed to validate this potentially useful biomarker.

## 1. Introduction

Huntington’s disease (HD) is an inherited neurodegenerative disease and hyperkinetic movement disorder involving the progressive loss of cognitive, behavioral, and motor abilities due an excess of CAG nucleotide repeats on the huntingtin gene. Currently there are no disease-modifying therapies for HD, and existing clinical predictors of symptom onset and progression have limited accuracy (Long et al. 2017). To develop reliable predictors and improve disease management strategies for this rare but debilitating and fatal condition, a better understanding of the biological mechanisms underlying HD progression is needed.

The predominate loss of medium spiny neurons in the striatum due to the presence of mutant huntingtin aggregates is thought to disrupt normal basal ganglia function and connectivity in HD [1], [2], [3]. In the cortico-basal ganglia-thalamo-cortical (CBTC) circuit, the striatum mediates indirect and direct signaling pathways that control function by receiving input signals from the cortex and regulating activity of the pallidum and substantia nigra pars reticulata. Downstream effects on thalamic function influence thalamo-cortical circuits that concern sensorimotor, cognitive, and limbic systems. The striatum also receives signaling from the intralaminar thalamic nuclei [4], [5], which indirectly facilitates information flow from the cerebellum. In the opposing direction, the striatum communicates with the cerebellum via intermediary subthalamic nuclei and brain stem pontine nuclei which in turn influence cerebellothalamic communication in a cyclic manner via the dentatorubrothalamic tract [6], [7]. Thus, any loss of striatal integrity threatening to imbalance CBTC circuit function could lead to engagement of thalamic and cerebellar nuclei to preserve function.

Emerging evidence supports the hypothesis that thalamic and cerebellar activity are altered in HD in a compensatory manner. For example, thalamocortical circuity involvement in early HD deficits has previously been suggested in electrophysiology studies of Q175 and R6/2 mice [8], [9]. This is further supported by earlier work demonstrating abnormal cortico-thalamic activations during impaired motor sequence learning in otherwise grossly asymptomatic patients with confirmed HD [10]. Striato-cerebellar hyper- and hypo-connectivity facilitated by the thalamus has also been reported in adolescent and teen gene carriers [11]. Pathologically, the cerebellum appears to be enlarged in patients with juvenile-onset HD despite striatal atrophy [12]. More recent work by our group has shown transient dentate nuclei enlargement with elevated susceptibility in adult premanifest HD, indicative of a compensatory change that could coincide with functional alterations [13]. Collectively, these data warrant further explicit investigation of thalamic and cerebellar functional alterations in HD, to both advance our understanding of the underlying disease mechanisms, as well as to identify candidate biomarkers of disease progression for clinical management and novel therapy trials.

The goal of this study was to use prospectively collected resting-state functional MRI (rsfMRI) data acquired at 7T, in addition to publicly available TRACK-HD data [14], to evaluate cross-sectional and longitudinal alterations in thalamo-cerebellar functional connectivity (FC) associated with HD progression.

## 2. Materials and Methods

### 2.1. Cohorts and Examinations

Participants in this study were prospectively recruited as part of a larger multimodal imaging study of HD (see Yao et. al. for full cohort details [13]). With local IRB approval, 21 patients with genetically confirmed HD (mean age 45, range 25-75; 48% female), and 13 healthy controls (mean age 40, range 26-69; 52% female) provided written informed consent to undergo tablet-based neurocognitive assessment [15], immediately followed by T1-weighted imaging and rsfMRI on a 7 Tesla (7T) MR950 research scanner (GE Healthcare, Milwaukee, WI, USA) with a 2-channel transmit and 32-channel receive coil. The neurocognitive tests and rsfMRI sequence parameters are detailed in **Table 1** (see Yao et. al. for other sequences acquired as part of the larger study protocol [13]). Specific cognitive tests were chosen to elucidate impairments in memory, executive function, visuospatial processing, and socioemotional aptitude that arise in the early disease-stage, often prior to motor symptom onset. To better characterize thalamo-cerebellar connectivity hypothesized to be altered on approach to symptom onset, we accessed 237 serial scans from 91 randomly selected premanifest HD patients (mean age 40, range 18-64) in the publicly available TRACK-HD dataset [14]. **Table 1** lists TRACK-HD rsfMRI parameters.

**Table 1.** Neurocognitive tests and MR parameters.

| Neurocognitive tests |  |
| --- | --- |
| Memory | <u>Favorites task</u> assessed verbal and visual associative memory. Subjects were asked to remember faces pairs with words describing their favorite food and animal. After 10-minutes, there was a delayed recall assessment, or <u>Favorites delay recall task</u>. The total number correct was scored for each memory test. |
| Executive Function | <u>Flanker task</u> assessed cognitive control and inhibitory attention. Subjects were shown five arrows and asked to quickly decide whether the center arrow was pointing left or right across congruent (e.g., all arrows same direction) and incongruent (e.g., center arrow opposite direction) trials. The equally weighed reaction time and accuracy was scored.<br><u>Set shifting task</u> assessed task switching abilities. Subjects had to match a visual stimulus to one of two other stimuli on the screen that involved either classifying shapes or colors in a homogeneous or pseudo-random fashion. The equally weighed reaction time and accuracy was scored.<br><u>Matching task</u> assessed executive function. Subjects were shown a legend of symbols, each with a unique number, and asked to select the symbol corresponding to the number(s) appearing in the middle of the screen. The total number correct was scored.<br><u>Timing task</u> assessed time perception. Subjects were visually presented with numerical measures of time and asked to start and stop a timer when they thought the presented time had passed. Their total error was scored. |
| Visuospatial Function | <u>Line orientation task</u> assessed visuospatial judgement. Subjects were shown three lines, a target line, and two other lines either parallel or non-parallel to the target line. They had to choose which line was parallel to the target line as difficulty increased. A threshold measure of accuracy was scored. |
| Socioemotional Function | <u>Dynamic affect recognition task</u> assessed emotional naming skills. Subjects were shown twelve 15-20s video clips of social interactions with semantically neutral scripts, and character emotion expressed through face expression, speech prosody, body language. They had to assign one of six possible emotions to the character and the total average reaction time was scored. |
| MRI parameters |  |
| T1-weighted | inversion recovery, spoiled gradient recalled sequence with inversion time=1350ms, repetition time (TR)=3.9ms, echo time (TE)=1.7ms, flip angle=8, 1mm isotropic voxel size, 25.6cm field-of-view (FOV), and in-plane ARC parallel imaging with acceleration factor of R=2.2 |
| fMRI (7T-HD) | interleaved, gradient echo (GRE) echo planar imaging (EPI) sequence with multiband factor 5, 291 time points, TR=1s, TE=22ms, flip angle=52, 1.6mm isotropic voxel size, 20.8cm FOV, and in-plane ARC parallel imaging with acceleration factor of R=2. |
| fMRI (3T-TRACK-HD) | T2* weighted echo planar imaging, 165 time points, TR = 3s, TE = 30 ms, flip angle = 80, 3.3x3.3x2.8mm voxel size, 21.2 mm FOV. |

### 2.2. HD Classification

Up to one month prior to 7T imaging, patients underwent neurological examinations with the Unified Huntington’s Disease Rating Scale (UHDRS [16]) to classify their disease stage based on their total motor score (TMS, range 0-124, lower score better), self-reported total functioning capacity (TFC, range 0-13, higher score better) and clinician diagnostic confidence level (DCL). HD gene carriers (CAG repeat ≥36) with TMS≤5, TFC=13, and a maximum DCL of 3, were classified as premanifest HD. Otherwise, patients were classified as manifest HD. To further characterize patient disease stage, a previously validated model [17] was used to estimate years to symptom onset (YTO) at the time of imaging from CAG repeats and age during the MRI. YTO estimates were also generated for TRACK-HD patients.

### 2.3. Data Processing

All MRI data were preprocessed and denoised using CONN, a MATLAB-based fMRI analysis toolbox [18]. The default preprocessing pipeline for volume-based analysis was used with default settings and included realignment, slice-timing, outlier-detection, segmentation, normalization to MNI space, and spatial smoothing using an 8mm kernel. These steps were preceded by removal of initial non-steady-state volumes (n=2), a step that was manually added to the pipeline. Denoising involved regression of default principal signal components derived from subject-specific white matter and cerebral spinal fluid segmentations, in addition to motion timeseries (n=12), and signal spikes flagged as outliers. To isolate neural signals of interest and maintain high degrees of freedom, the data were finally bandpass filtered between 0.008 and 0.15Hz. Average fMRI signal within each thalamic and cerebellar region-of-interest (ROI) were automatically calculated by CONN (e.g. ROI_Subject001_Condition001.mat in results directory) using their default brain atlas, a combination of the Harvard-Oxford and AAL atlases. A second longitudinal deep gray matter atlas derived from quantitative susceptibility mapping data [19] was manually imported into CONN such that averaged fMRI signal of the cerebellar dentate nucleus–previously found to be enlarged in premanifest HD [13]–could also be extracted. For each left and right cerebellar ROI (anterior, posterior, and flocculonodular lobes, and the dentate nucleus), connectivity to the ipsilateral and contralateral thalamus was calculated in MATLAB (MathWorks, Natick, Massachusetts) as the Fisher-transformed Pearson correlation between ROI timeseries. This approach yielded a total of 16 connectivity measures per subject.

### 2.4. Analysis

The overarching goal of the analysis was to identify thalamo-cerebellar FC markers of HD progression and compensation using a priori knowledge of thalamic function and its structural connections to the cerebellum. Analyses were performed in MATLAB, solely using clinical metrics and connectivity measures computed as described in section 2.3.

#### 2.4.1. 7T-HD cohort: controls, premanifest, and manifest HD

Building upon our prior finding of a transient increase in volume and iron deposition in the dentate nucleus in premanifest HD [13], we first sought to evaluate cross-sectional differences in FC across healthy controls, premanifest HD, and manifest HD. Multiple Kruskal-Wallis tests were run with post hoc Dunn’s multiple comparisons tests and effect sizes calculated as η^2^, the Kruskal-Wallis chi-square statistic divided by N-1, where N is the total number of subjects across all groups. To further investigate group-level effects, the relationship between FC strength of significant ROIs (p_Kruskal_<0.05) and TMS, as well as estimated YTO, was assessed using Pearson correlations and polynomial fitting with a significance level of 0.05. Bonferroni-adjusted p-values were also computed to determine significance under multiple comparison. Finally, group differences in cognitive performance were evaluated using the same statistical tests described above; cognitive scores yielding significant group differences were combined into a composite score (task-specific z-score average) that was thereafter correlated with the most relevant thalamo-cerebellar connections, based on their relationship to disease severity.

#### 2.4.2. TRACK-HD cohort: premanifest HD

To more narrowly investigate FC patterns observed in the 7T-HD cohort and test the hypothesis that premanifest HD is associated with transient brain alterations, TRACK-HD subjects were first separated into five groups based on their TMS (0, 1, 2, 3 or 4), excluding two individuals with a TMS of 5. TRACK-HD group differences in FC were cross-sectionally evaluated using Kruskal-Wallis tests, and FC was subsequently related to YTO via Pearson correlations (a primary significance level of 0.05 was used for all tests, followed by Bonferroni adjustment). Based on findings in the 7T-HD cohort, we opted to evaluate patient’s left and right FC values together as independent measures, keeping ipsilateral and contralateral FC values separate. An exploratory longitudinal analysis followed for a subset of patients with suitable serial data, specifically three similarly spaced imaging time points and YTO ranging 0 and 15 years (the anticipated period of transient alterations and most dense range of patient YTO values). The change in FC was calculated across time points 1 and 3 and then correlated with YTO. Non-parametric regression via locally weighted scatterplot smoothing (lowess) was then applied to the graphs using an experimental smoothing factor of 0.6, to explore patterns in the data without any assumption about the specific distribution they may follow. Here, left and right FC values were also mapped together while ipsilateral and contralateral FC values remained separate.

## 3. Results

### 3.1. Observations

Subject age, sex, education, and other clinical parameters at the primary imaging time point are summarized in **Table 2**. Of the 91 TRACK-HD subjects, 42 had suitable serial imaging data that was included in the longitudinal analysis (see 2.4.2) with median follow up times of 351 days (range 251-483) for time point 2 and 709 days (range 566-882) for time point 3. As shown in **Figure 1**, YTO significantly (p<0.05) correlated with CAG for both the 7T-HD and TRACK-HD groups (r_7T_=-0.56, r_TRACK_=-0.25), as well as TFC scores (r_7T_=0.48) for the 7T-HD group. Based on this result, we proceeded to use YTO in the analyses to further distinguish patients beyond their discrete TMS scores.

**Figure 1.**
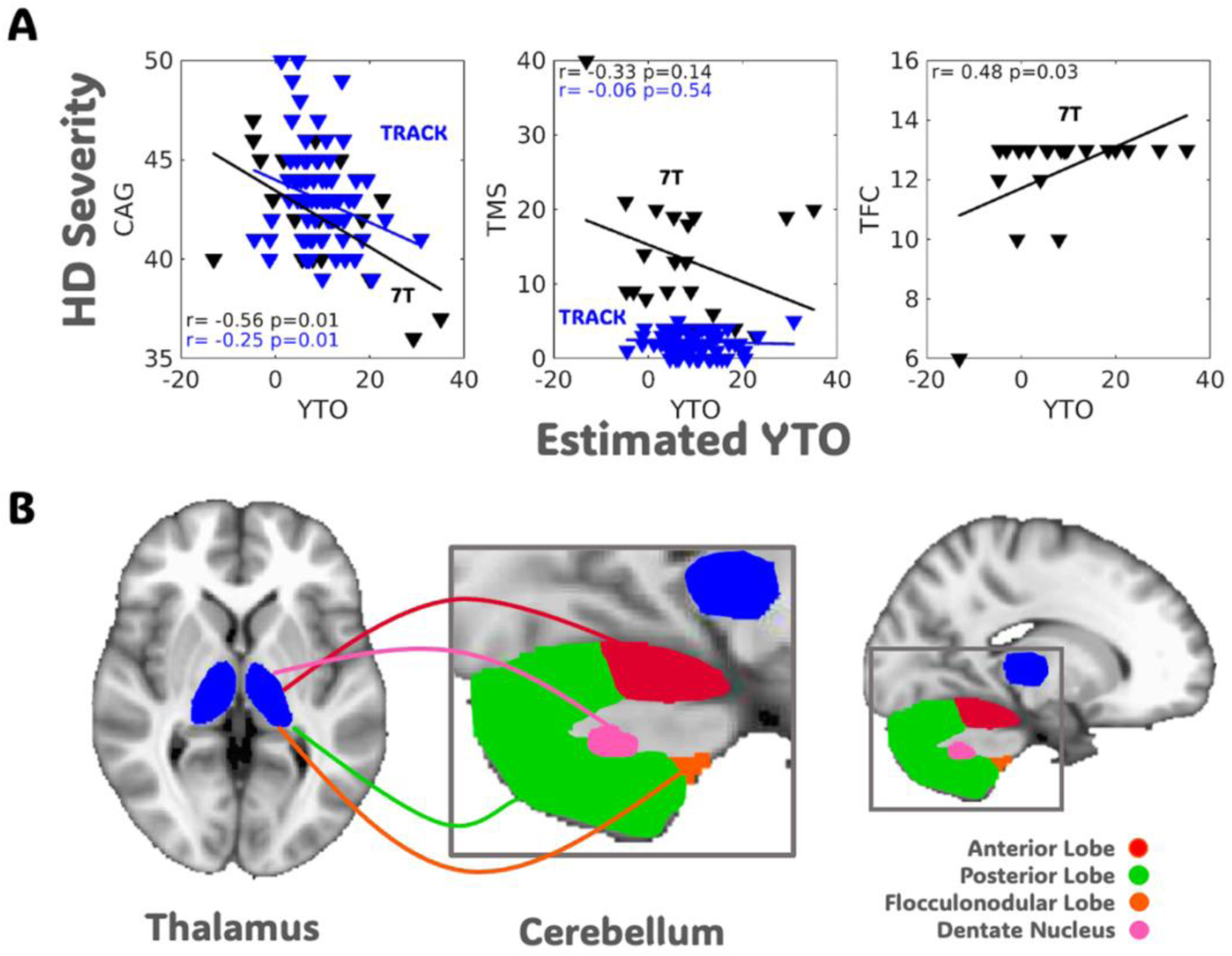
HD severity vs. estimated years to symptom onset and brain regions of interest. **A.** All data are from the first imaging timepoints. Years to symptom onset (YTO) significantly correlated with CAG repeats for both 7T-HD and TRACK-HD subjects, as well as available total functioning capacity (TFC) scores for the 7T-HD group. **B.** Connectivity between the thalamus and each cerebellar lobe, as well as the dentate nucleus, were the compared across patient groups and correlated with YTO and total motor score (TMS).

**Table 2.** Cross-sectional patient characteristics.

| Characteristic | Subject Proportions |  |  |  |  |
| --- | --- | --- | --- | --- | --- |
|  | N<sub>HC</sub>=13 | N<sub>HD</sub>=21 | N<sub>PM</sub>=4 | N<sub>M</sub>=17 | N<sub>TRACK-HD</sub> = 91 |
| Age, mean(range) | 40(26-69) | 45(25-75) | 38(25-49) | 48(27-75) | 40(18-64) |
| Female, (%) | 52 | 48 | 50 | 47 | - |
| Race/Ethnicity, (%) |  |  |  |  |  |
| white | 7 | 100 | 100 | 100 | - |
| asian | 4 | - | - | - | - |
| hispanic | 1 | - | - | - | - |
| unknown | 1 | - | - | - | - |
| Years of education, mean(range) | 18(16-25)<sup>a</sup> | 16(12-21)<sup>a</sup> | 15(14-16)<sup>a</sup> | 16(12-21)<sup>a</sup> | - |
| CAG repeats, median(range) | - | 42(36-47) | 42(39-43) | 43(36-47) | 43(39-50) |
| UHDRS-TMS, median(range) | - | 13(1-40) | 4(1-4) | 14(6-40) | 2(0-5) |
| UHDRS-TFC, median(range) | - | 13(6-13) | 13(13-13) | 13(6-13) | - |
| predicted YTO, median(range) | - | 8(-13-35) | 19(9-23) | 5(-13-35) | 8(-4-30) |

<sup>a</sup>education unavailable for 3HC, 2PM, 5M

### 3.2. 7T-HD cohort: controls, premanifest, and manifest HD

#### Group differences

Significant group differences in thalamic and dentate nucleus FC (**Figure 2a**, **Table 3**, p_kruskal_<0.05) were observed for both ipsilateral and contralateral connections of the right thalamus (premanifest vs. control: p_Dunn_<0.05, η^2^_ipsilateral_=0.22, η^2^_contrlateral_=0.19), and the contralateral connection of the left thalamus (premanifest vs. control: p_Dunn_<0.05, η^2^=0.20). FC was visibly reduced in premanifest HD subjects, resembling an inverted arc-like pattern, and post hoc testing indeed revealed significant pair-wise FC differences across premanifest HD and healthy controls. However, no tests surpassed the Bonferroni-adjusted significance threshold (p=0.003, N_tests_=16).

**Figure 2.**
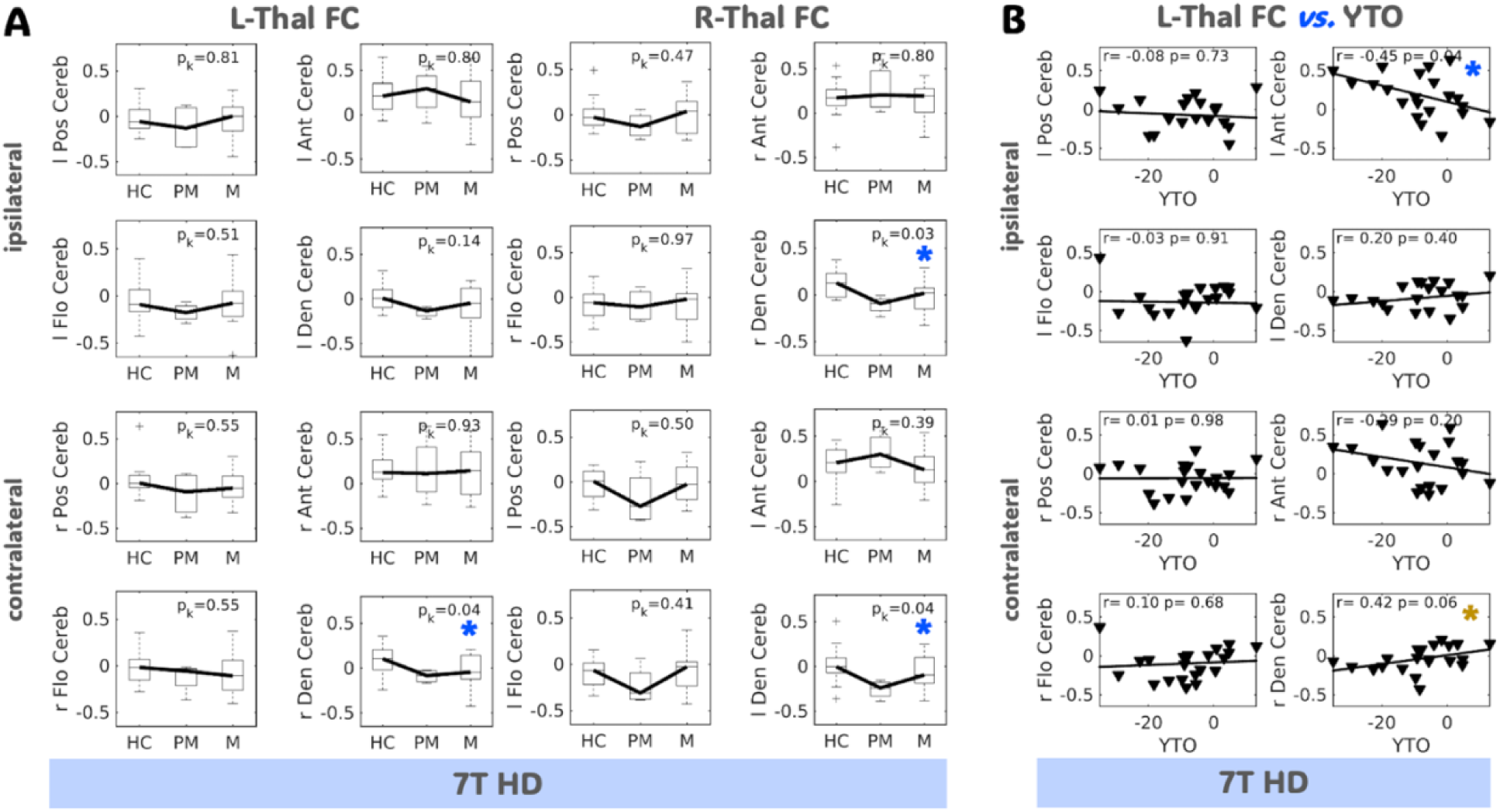
7T HD group differences in thalamo-cerebellar connectivity and relationship to years to onset. **A.** Significant group differences were observed for ipsilateral and contralateral thalamic (Thal) connections to the dentate nucleus (Den Cereb), as denoted by the blue stars. **B.** Patients with fewer estimated years to onset (YTO; made negative here to illustrate *decreasing* years to onset from *left to right*) had significantly decreased thalamic functional connectivity (FC) to the anterior cerebellar lobe (Ant Cereb). The opposite (near-significant) trend was seen for the deep gray matter Den Cereb: FC with the thalamus increased with decreasing YTO as denoted by the gold star. HD=Huntington’s disease; HC=health control; PM=premanifest; M=manifest; l=left; r=right; Ant Cereb=anterior lobe; Flo Cereb=flocculonodular lobe

**Table 3.**
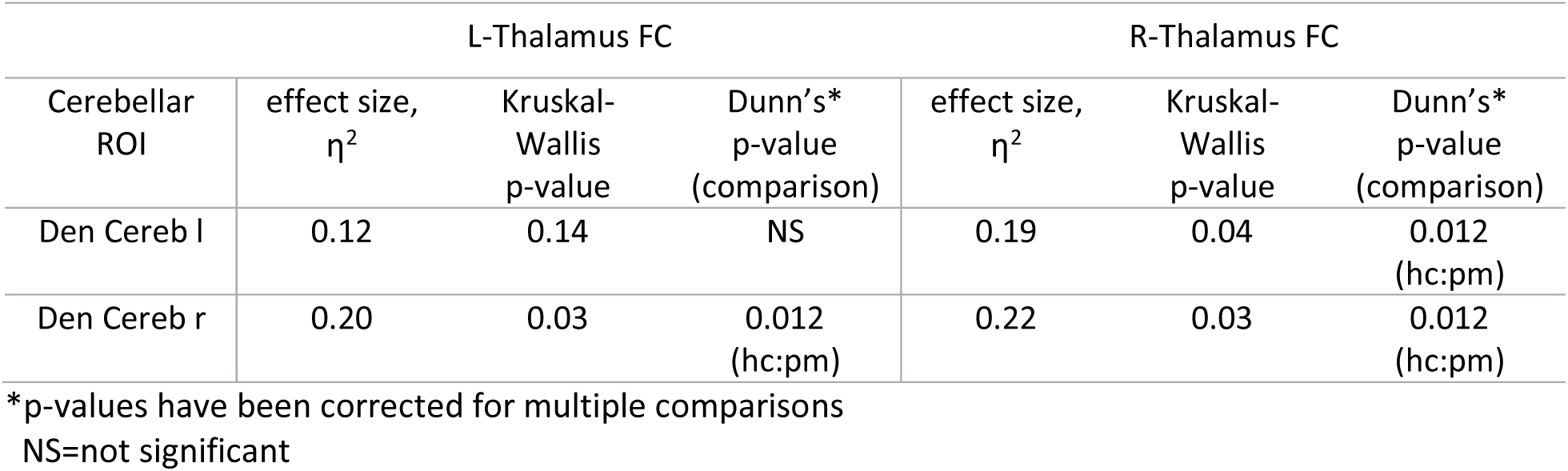
7T-HD group differences in connectivity.

| Cerebellar ROI | L-Thalamus FC |  |  | R-Thalamus FC |  |  |
| --- | --- | --- | --- | --- | --- | --- |
| | effect size, $\eta^2$ | Kruskal-Wallis p-value | Dunn's* p-value (comparison) | effect size, $\eta^2$ | Kruskal-Wallis p-value | Dunn's* p-value (comparison) |
| Den Cereb l | 0.12 | 0.14 | NS | 0.19 | 0.04 | 0.012 (hc:pm) |
| Den Cereb r | 0.20 | 0.03 | 0.012 (hc:pm) | 0.22 | 0.03 | 0.012 (hc:pm) |

\*p-values have been corrected for multiple comparisons
NS=not significant

#### Relationship with YTO and TMS

When correlating FC and YTO across patients, left thalamic FC to the ipsilateral anterior cerebellum was found to significantly decrease the closer an individual was to symptom onset (**Figure 2b**, r=-0.45, p<0.05); this was consistent with a predicted decline in FC following a transient increase. The opposite pattern was observed for left thalamic FC to the contralateral dentate nucleus; this was a near-significant effect (r=0.42, p=0.06). A significant positive correlation was furthermore observed between YTO and the right thalamic FC to the ipsilateral dentate nucleus (**Supplementary Figure 1**). Finally, plotting FC against TMS also revealed similar decreasing and increasing patterns for thalamic connections to the anterior cerebellum and dentate nucleus, however, they were not significant (**Supplementary Figure 2**). Again, no tests for this small cohort surpassed the Bonferroni-adjusted significance threshold (p_adjust_<0.003, N_tests_=16).

#### Relationship with cognitive performance

Significant group differences in cognitive performance (**Figure 3a**, **Table 4**, p_kruskal_<0.05) were observed for the following four domains: inhibition (manifest vs. control: p_Dunn_<0.01, η^2^=0.28), task switching (manifest vs. control: p_Dunn_<0.01, η^2^=0.30), executive function (manifest vs. control: p_Dunn_<0.01, η^2^=0.29), and visuospatial judgement (manifest vs. premanifest: p_Dunn_<0.01, η^2^=0.24). Near-significant group differences were also seen for the delayed memory recall task; none of the comparisons surpassed the Bonferroni-adjusted threshold. As shown in **Figure 3b**, a composite measure (task-specific z-score average) of the four significant cognitive tasks correlated with thalamic FC to the ipsilateral and contralateral anterior cerebellum (r=0.18, p<0.05) and dentate nucleus (r=-0.17, p<0.05) across all subjects. Like the relationship observed between FC and YTO (**Figure 2b**), worse performance coincided with decreased thalamo-anterior cerebellar FC and increased thalamo-dentate nucleus FC. These effects were persistent and strong when evaluating groups separately (**Supplementary Figure 3**): controls demonstrated significant anterior cerebellar effects (r=0.63, p<0.0001), and patients significant dentate nucleus effects (r=-0.43, p<0.0001), both of which were significant after Bonferroni correction (p_adjust_<0.01, N_tests_=4).

**Figure 3.**
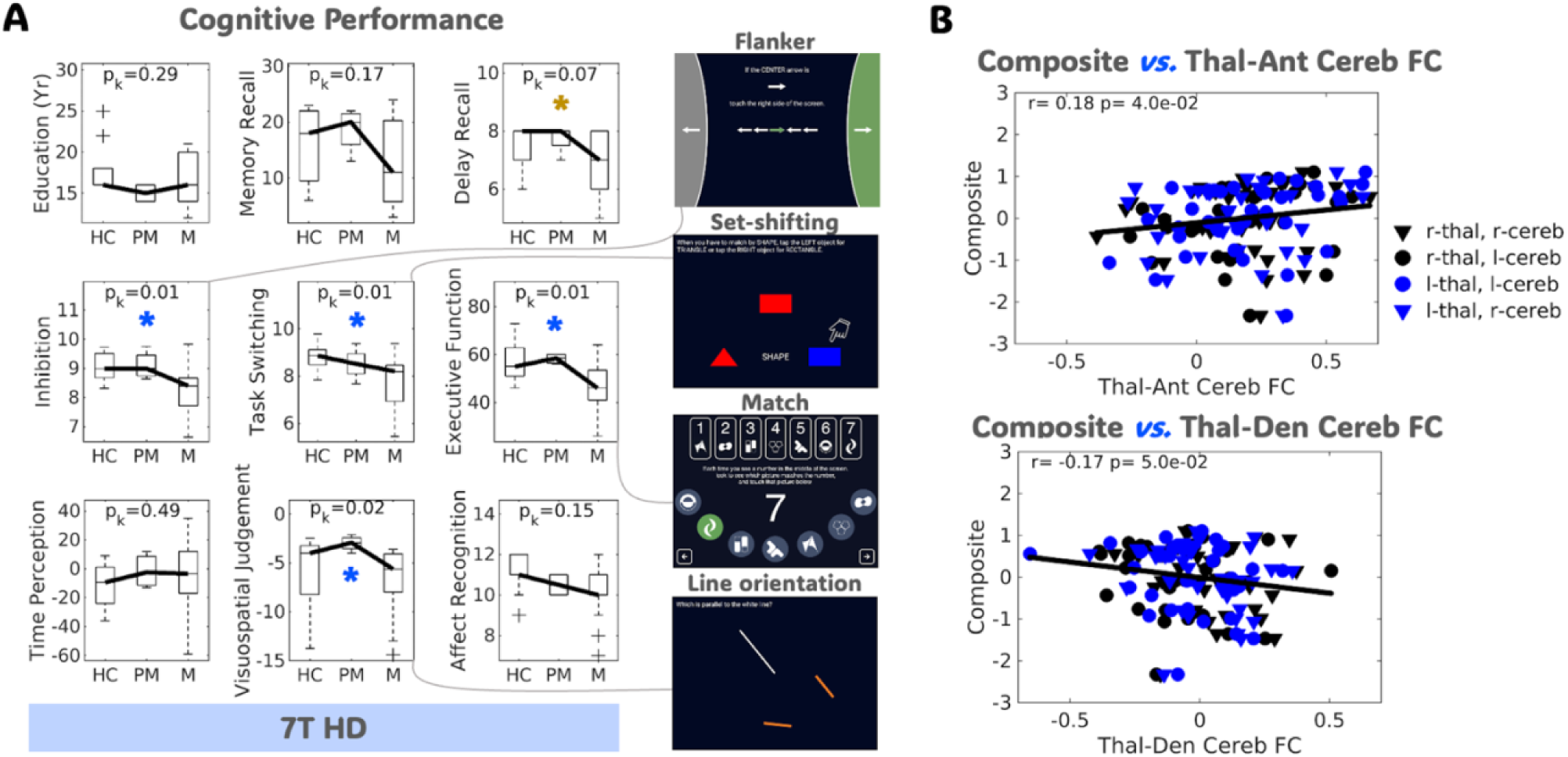
7T HD group differences in cognitive performance and relationship to connectivity. **A.** Significant group differences, primarily between health control (HC) and manifest (M) patients, were observed for cognitive tests involving inhibition (Flanker), task switching (Set-shifting), executive function (Match), and visuospatial judgement (Line orientation), as denoted by the blue stars. Memory recall (Favorites) also showed near-significant group differences as denoted by the gold star. **B.** A composite measure of cognitive performance derived from tests with significant group differences revealed significantly lower (worse) cognitive performance with lower functional connectivity (FC) between the thalamus (Thal) and anterior cerebellar lobe (Ant Cereb); the opposite effect was observed for thalamic FC to the dentate nucleus (Den Cereb). HD=Huntington’s disease; PM=premanifest; l=left; r=right

**Table 4.** 7T-HD group differences in cognitive performance.

| Domain | Cognitive performance |  |  |
| --- | --- | --- | --- |
| | effect size, $\eta^2$ | Kruskal-Wallis p-value | Dunn's* p-value (comparison) |
| Education (yrs) | 0.08 | 0.29 | NS |
| Memory Recall | 0.12 | 0.17 | NS |
| Delay Recall | 0.16 | 0.07 | NS |
| Inhibition | 0.28 | 0.01 | 0.007 (hc:m) |
| Task Switching | 0.30 | 0.01 | 0.002 (hc:m) |
| Executive Function | 0.29 | 0.01 | 0.008 (hc:m) |
| Time Perception | 0.04 | 0.49 | NS |
| Visuospatial Judgement | 0.24 | 0.02 | 0.006 (pm:m) |
| Affect Recognition | 0.12 | 0.14 | NS |

\*p-values have been corrected for multiple comparisons
NS=not significant

### 3.3. TRACK-HD cohort: premanifest HD

#### Group differences and relationship with YTO

Whereas cognitive effects were primarily limited to differences in performance between patients with manifest HD and controls (**Figure 3**), FC effects were driven by premanifest HD and control group differences in connection strength. Further investigating the latter, patterns of transient increase (or decrease) in FC were observed for TMS scores ranging 1-2 (**Supplementary Figure 4**), and though not significant, plotting ipsilateral FC for each of the five patient groups (TMS 0-4) against YTO revealed significant effects (**Figure 4**): FC increased with decreasing YTO for lower TMS groups (r=0.56-0.64, p<0.01-p<0.001) and decreased with decreasing YTO for higher TMS groups (r=-0.29-0.49, p<0.05-p<0.001) consistent with a predicted transient change in FC. A similar observation was made for contralateral FC comparisons (**Supplementary Figure 5**) and several comparisons relating thalamo-posterior cerebellar FC to YTO surpassed the Bonferroni-adjusted significance threshold (p_adjust_<0.001, N_tests_=40). Importantly, some comparisons (0>TMS<4) yielded Pearson correlations near zero, which was deemed significant here based on the hypothesis that transitory changes in FC could be chronologically defined as an initial change, then stabilization, then reversion to baseline.

**Figure 4.**
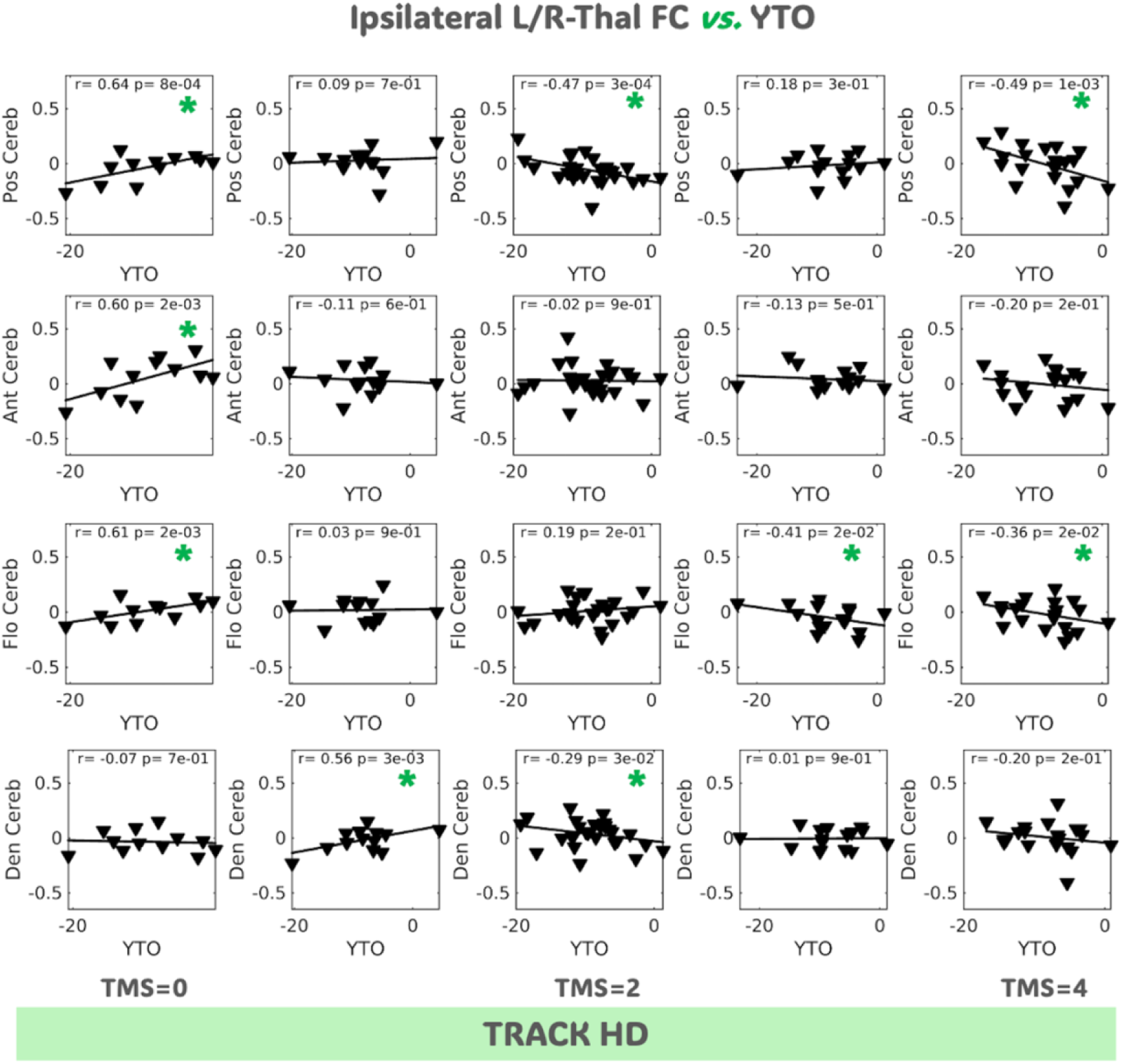
Relationship between ipsilateral thalamo-cerebellar connectivity and years to onset in TRACK-HD cohort. Patients were separated into five groups based on their total motor scores (TMS) ranging 0 to 4. For individuals with low TMS (0-1), multiple thalamo-cerebellar connections showed significantly increased functional connectivity (FC) with decreasing estimated years to onset (YTO; made negative here to illustrate *decreasing* years to onset from *left to right*) as denoted by the green star. The opposite effect was seen for individuals in the high TMS (2-4) group. Thal=thalamus; HD=Huntington’s disease; l=left; r=right; Pos Cereb=posterior lobe; Ant Cereb=anterior lobe; Flo Cereb=flocculonodular lobe; Den Cereb=dentate nucleus

#### Exploratory longitudinal analysis

Individual FC trajectories varied over time (example shown in **Figure 5a**): some patients’ FC increased, remained static, or decreased. Non-parametric regression via lowess (see 2.4.2.) revealed the predicted arc-like pattern indicative of transient FC change, whereby patient-level thalamo-anterior cerebellar FC change plotted across all subjects progressed from negative to positive, then negative again, with decreasing years to onset. Lowess fits were more linear for posterior and flocculonodular lobe FC, with a subtle inverted arc for dentate nucleus FC that was mostly consistent with cross-sectional findings.

**Figure 5.**
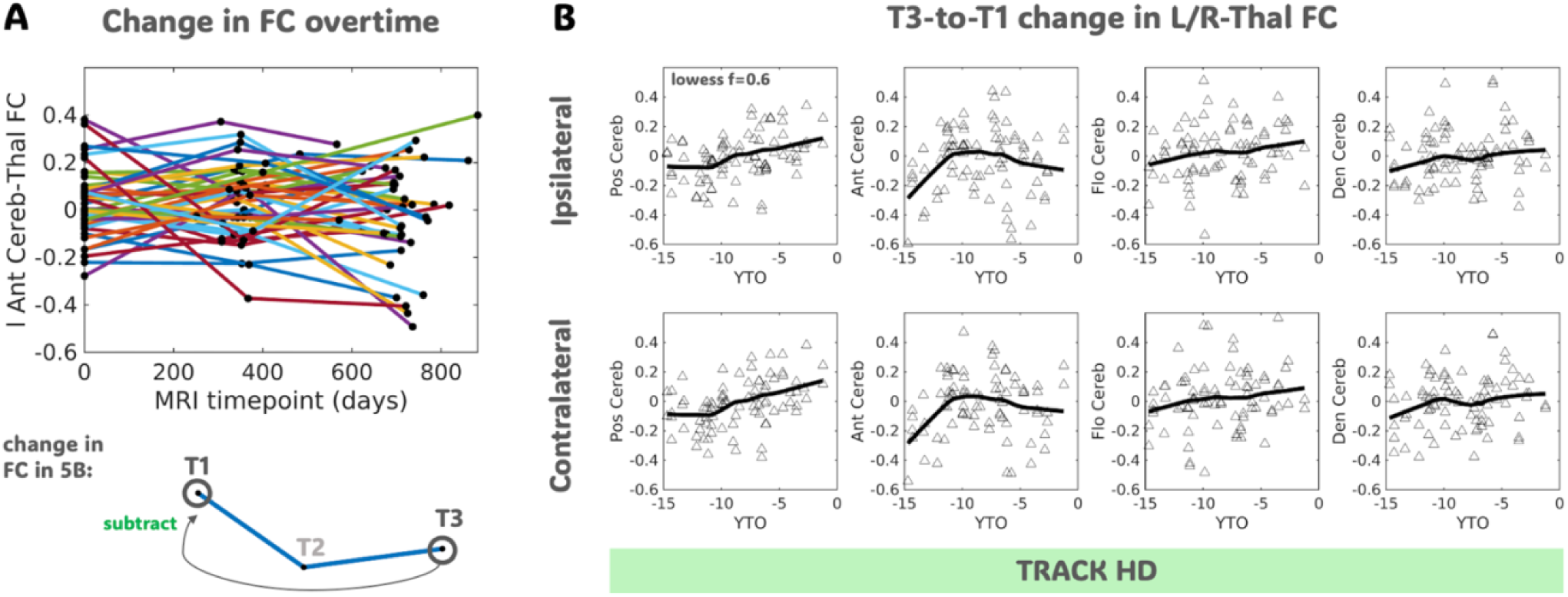
Longitudinal patterns in TRACK-HD thalamo-cerebellar connectivity with years to onset. **A.** Forty-two patients showed changes in thalamo-cerebellar functional connectivity (FC) over time; a representative group example is shown for the left ipsilateral thalamic (Thal) connection to the anterior lobe (Ant Cereb). Change in FC was calculated by subtracting time point 3 (T3) from time point 1 (T1). **B.** Non-parametric regression using locally weighted scatterplot smoothing (lowess) with a smoothing factor of 0.6, revealed linear and parabolic patterns in FC with decreasing estimated years to onset (YTO; made negative here to illustrate *decreasing* years to onset from *left to right*) similar to cross-sectional findings. Pos Cereb=posterior lobe; Flo Cereb=flocculonodular lobe; Den Cereb=dentate nucleus

## 4. Discussion

This study aimed to investigate thalamo-cerebellar connectivity in HD and further our understanding of potential biological mechanisms underlying HD progression. Our main finding was that there appeared to be transient changes in thalamo-cerebellar connectivity during the premanifest phase of HD. In the smaller 7T group of healthy controls and patients with HD, this transient effect was predominately evident for thalamic connections to the anterior lobe and dentate nucleus. In the larger TRACK-HD group of premanifest HD, these pattern extended to thalamic connections to the posterior and flocculonodular lobes, with serial data and non-parametric regression providing additional promising evidence of transient change in thalamo-anterior cerebellar FC.

There are several pieces of evidence and hypotheses in the literature to potentially explain these findings. First, our group recently performed ROI-based volumetric analysis and found that the dentate nuclei are significantly enlarged in premanifest HD and have more iron as evidenced by increased susceptibility relative to both healthy controls and patients with manifest HD [13]. Prompted by this finding, we evaluated connectivity in a subset of patients from the same cohort who therefore now have evidence of both transient volume and connectivity change involving the cerebellum. While several voxel-based morphometry studies have reported normal cerebellar volume in premanifest HD [20], [21], one group reported cerebellar enlargement in juvenile-onset HD patients [12]. In more advanced disease stages, postmortem studies have reported cerebellar degeneration at a rate consistent with symptom severity [22], [23]. Taken together, it is plausible that enlargement of the cerebellum could reflect a transient process in HD as part of a larger compensatory mechanism that has been hypothesized in the literature [24].

To explain the morphologic phenomenon of cerebellar enlargement in HD and relate it to transient changes in functional connectivity, one might consider the demyelination hypothesis in HD [25], [26]. This evidence-based hypothesis proposes the premature breakdown of myelin leading to a surge in production of iron-rich oligodendrocytes for repair [27], [28], [29]. Ultimately, oligodendrocyte production is thought to be halted by iron toxicity and/or glial cell dysfunction, explaining the lack of these cells in patients at the symptomatic HD stage [30]. Accumulation of oligodendrocytes can alter apparent cortical boundaries on imaging, directly influencing gray matter volume measurements [31], and could be accompanied by transient alterations in connectivity as myelin integrity plays a key role in facilitating and maintaining synchronized transfer of neural information [32]. More specifically, the notion of transient connectivity *increase* due to premature myelin breakdown aligns best with the arc-like pattern in connectivity we observed for thalamo-anterior cerebellar connections, as well as prior evidence of compensation through connectivity increase across the lifespan and in neurodegenerative disease [33], [34]. Additional strong evidence supporting this comes from a metabolic imaging study which reported hypermetabolism of the ventrolateral thalamus and cerebellar vermis in the preclinical disease stage up until symptom onset at which point metabolic activity fell to subnormal levels [35]. Notably, in the 7T-HD cohort, dentate nucleus connectivity to the thalamus showed the opposite pattern (an inverted arc-like pattern) to that of the anterior cerebellum. Both regions have known involvement in complex sensorimotor functions [36], [37], but the dentate nucleus previously found to show transient increase in volume and iron deposition in premanifest HD [13], was associated with reductions in connectivity with progression. One hypothesis is that these potential pathologic alterations the dentate nucleus is undergoing, coincide with local reductions in functional integrity and thus dentate connection strength to the thalamus, which may in turn impact thalamic connectivity with the cerebellar cortex (such as the anterior lobe) to which the dentate nucleus is structurally and functionally connected [38].

Adding to these findings, results from TRACK-HD group suggest that patients with low TMS ≤1 are experiencing increased thalamic connectivity to multiple cerebellar regions, the closer they are to estimated symptom onset (a prediction based on their CAG repeats and age). Although we did not have ample data with YTO>20 to characterize very early brain connectivity, our 7T-HD data suggest that normal thalamo-cerebellar connections are either weaker or stronger than that of premanifest HD depending on the cerebellar ROI. In support of the latter, a recent study of adolescent and teen gene carriers reported an early decline in striato-cerebellar connectivity, mediated by the thalamus, from a hyper- to hypo-connected state with increasing age and approach to disease onset [11]. Though not explicitly measured within the thalamus or cerebellum, reduced sensorimotor connectivity in premanifest HD has also been found relative to healthy controls [39].

On the opposite end, in the late premanifest to manifest disease stage, cross-sectional TRACK-HD and 7T-HD data suggest that connectivity returns to a relative baseline state. However, this was contrasted by some of the exploratory longitudinal results for specific cerebellar ROIs like the posterior lobe that show subtle FC increases with decreasing YTO (similar to that of the 7T-HD cohort) and not the predicted arc-like trend observed for thalamo-anterior cerebellar connections. One potential source of variability that may explain these differences, is the joint evaluation of patients with variable TMS scores in the longitudinal analysis to maintain a sufficient sample size for lowess regression. Nonetheless, given the progressive nature of HD and postmortem evidence of cerebellar and more widespread atrophy [22], [23], [40], its plausible that connectivity could continue on a worsening trajectory, be it increased and/or decreased FC between the thalamus and cerebellar subregions. However, this remains to be investigated in a larger dataset with more dense sampling across early and late HD stages.

While a thorough evaluation of cognitive performance was also not the focus of this work, it provided additional evidence to strengthen our hypothesis that transient alterations in thalamo-cerebellar connectivity could be related to compensatory mechanisms in HD. In several domains, premanifest patients from the 7T-HD cohort performed as well as healthy controls, or slightly worse, despite evidence of significant differences in connectivity. Similar findings have been reported for the TRACK-HD cohort in a multimodal analysis relating global cognitive performance with functional alterations and structural disease load [41]; other findings from the PREDICT-HD study have also linked cognition to dynamic brain alterations [42]. Interestingly, thalamo-anterior cerebellar connectivity of healthy controls alone significantly correlated with composite cognitive performance scores. This could reflect reaction time and thus motor planning [43], as reaction time was a primary or factored measure of performance for several cognitive tests included in the composite scores; though it may also reflect growing evidence of cerebellar involvement in multiple cognitive processes [44], [45].

Finally, this study has several limitations. Evaluating connectivity of the whole thalamus and cerebellar lobes as opposed to their constituent nuclei and lobules mitigates noise, but also limits interpretation of the biological significance of our results. The prospective 7T-HD cohort is also small and heterogeneous, in part due to HD being a rare disease and because fMRI data was only collected for a subset of the larger study cohort [13]. To balance this, we included a larger and more uniform TRACK-HD cohort. Nonetheless, the limited number of premanifest patients in the 7T cohort along with the grouping of all symptomatic patients as manifest HD reduces reliability of the results. The low sample size and limited statistical power, especially for the 7T-HD cohort, prompted us to integrate exploratory regression techniques like lowess and furthermore treat left and right FC as independent measures, combining them in some analyses. For this reason, we could not confidently state the clinical significance of our results, though we do believe this work supports continued investigation of thalamo-cerebellar connectivity as a candidate biomarker of HD progression. Last, we did not thoroughly investigate cognitive performance in the TRACK-HD cohort as this was beyond the scope of this work; TRACK-HD data were incorporated solely to resolve connectivity patterns observed in premanifest HD, and in fact most significant group differences in cognitive performance did not involve premanifest HD, but manifest HD and controls. It is also important to note that YTO is a predicted measure and should be interpreted as such. Here we used YTO to differentiate patients beyond their TMS, especially in the premanifest stage where multiple patients shared the same score. In demonstrating that YTO correlates with various measures of disease severity, the assumption is that the metric can provide a useful estimate of disease stage; however, it may not be the most optimal approach for clustering patients given that YTO can vary significantly for patients with the same motor score. Coupled with the fact that the TMS is a subjective clinical measure that can vary by day and rater, the result should be carefully interpreted. Nonetheless, despite these limitations, the results align with growing evidence of aberrant connectivity in HD involving cerebellar and thalamic connections, providing new evidence of transiently modulated connectivity between these two structures, with progression.

## 5. Conclusion

In conclusion, our multi-institutional functional imaging study provides some evidence of transient alteration in thalamo-cerebellar connectivity in asymptomatic premanifest HD. This work contributes complimentary information to a growing body of literature, detailing compensatory mechanisms underlying HD progression. Promising biomarkers for patient stratification could emerge with larger-scale quantitative investigations of these thalamo-cerebellar connections that undoubtingly play a role in HD manifestation.

## Data Availability

All UCSF data used in the present study are available upon reasonable request to the authors. All TRACK-HD data were made available by CHDI Foundation, Inc.

## Acknowledgements

The authors would like to acknowledge the support of MRI staff in the Surbeck Laboratory for Advancing Imaging at UCSF, and General Electric Healthcare for their support with 7T sequence development. The authors also thank the Memory and Aging Center at UCSF, and patients and caregivers for their participation in this study. Data and data derived from biosamples used in this work were generously provided by the participants in the TRACK-HD study and made available by Professor Sarah Tabrizi, Principal Investigator, University College London and Dr. Edward Wild, University College London.

## Supplementary Figures

**Supplementary Figure 1.**
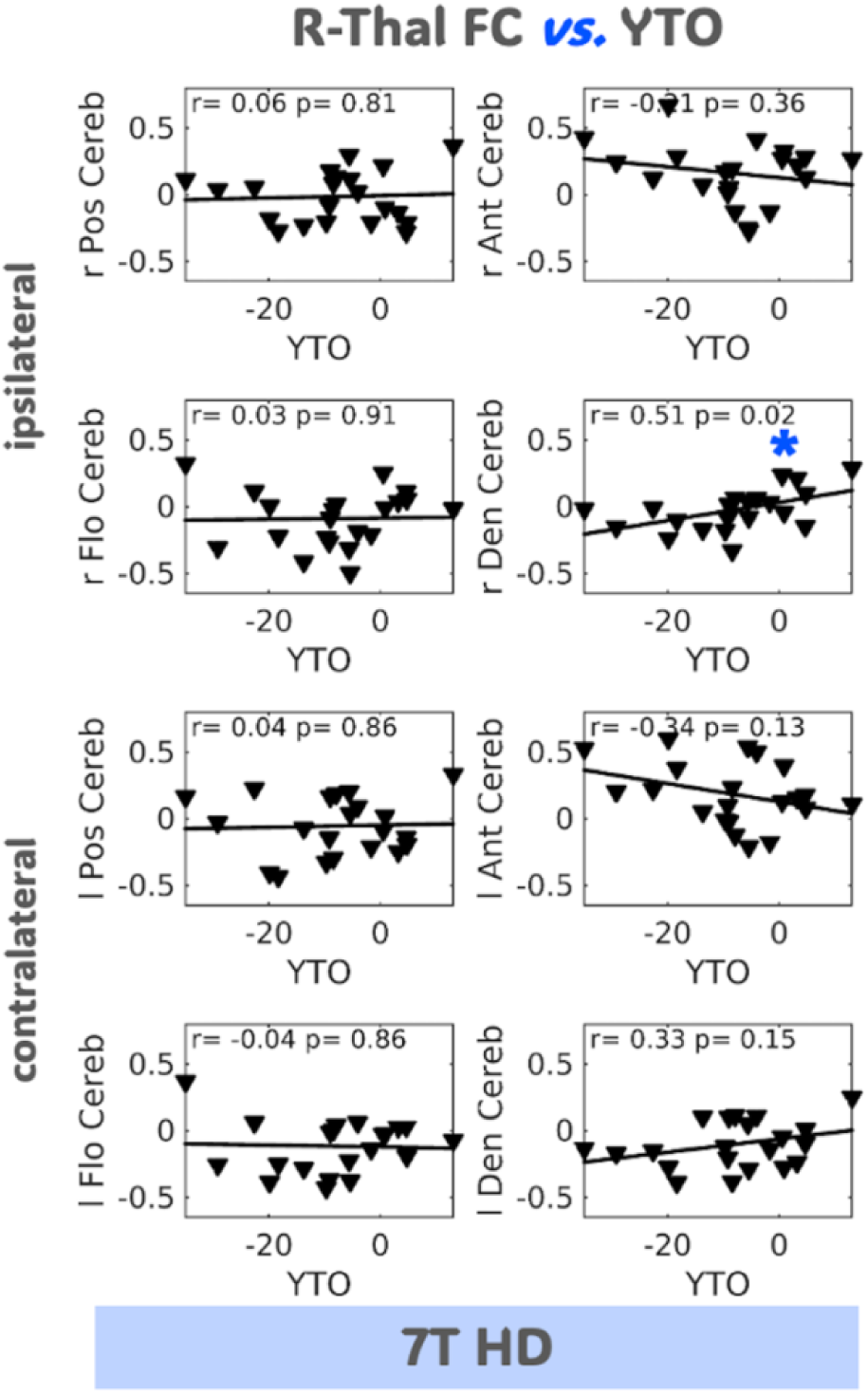
Relationship between thalamo-cerebellar connectivity and years to onset in 7T HD cohort. As shown for the left (l) thalamus (Thal) in Figure 2B of the main text, functional connectivity (FC) between the right (r) thalamus and dentate nucleus (Den Cereb), significantly increased with *decreasing* years to onset (YTO; made negative here to illustrate *decreasing* years to onset from *left to right*) as denoted by the blue star. Although not significant, r-thalamic FC to both the ipsilateral and contralateral anterior cerebellar lobes (Ant Cereb), showed the opposite trend, mirroring that observed for the l-thalamus in Figure 2B of the main text. HD=Huntington’s disease; YTO=years to onset; Pos Cereb=posterior lobe; Flo Cereb=flocculonodular lobe

**Supplementary Figure 2.**
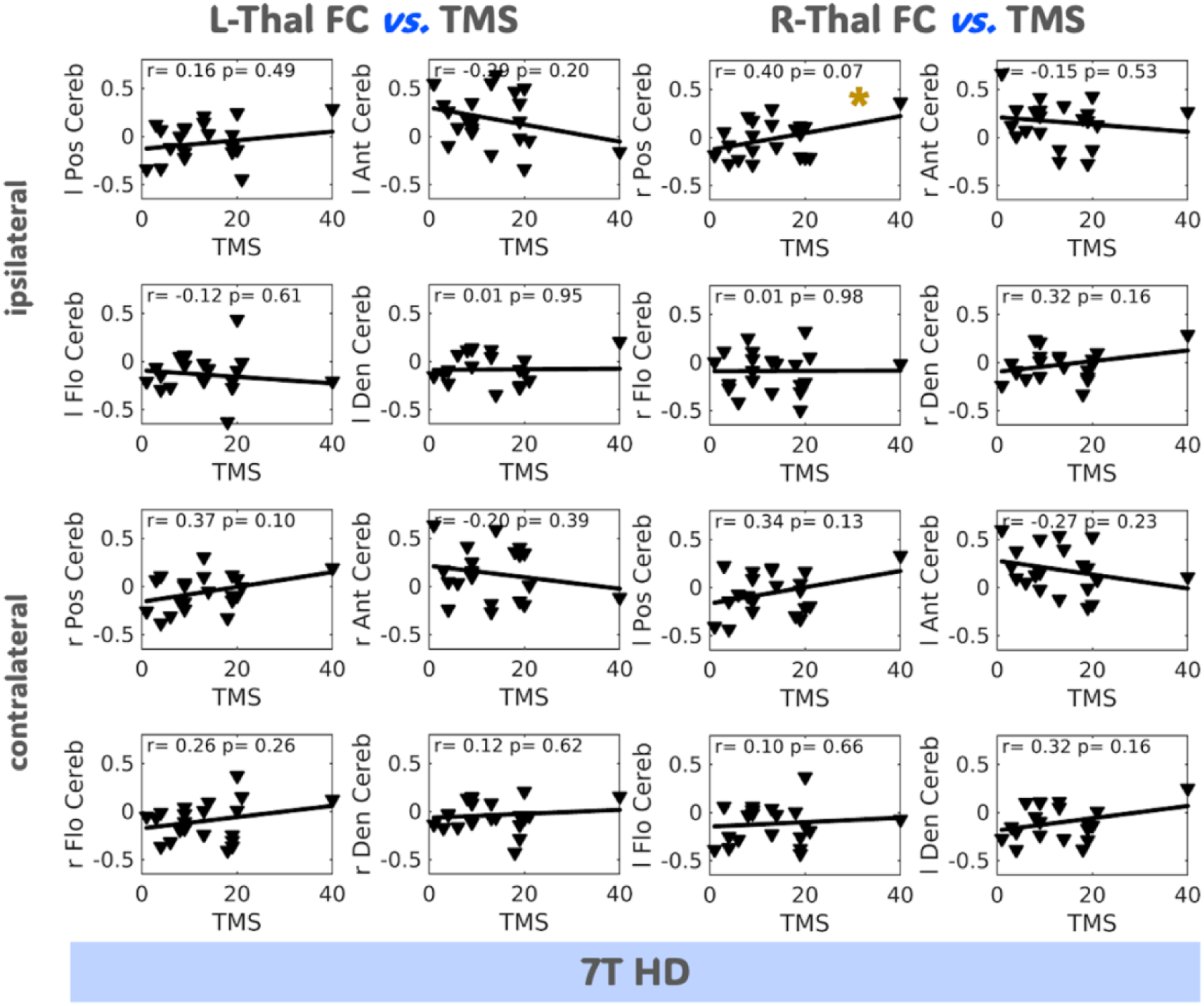
Relationship between thalamo-cerebellar connectivity and total motor score in 7T HD cohort. Although not significant, a trend of increasing thalamic-cerebellar functional connectivity (FC) with increasing disease severity was observed for the posterior cerebellar lobe (Pos Cereb; denoted by the gold star) and some dentate nucleus (Den Cereb) connections. The opposite trend was observed for the anterior cerebellar lobe (Ant Cereb): FC with the thalamus decreased with increasing disease severity. HD=Huntington’s disease; TMS=total motor score; l= left; r= right; Flo Cereb=flocculonodular lobe

**Supplementary Figure 3.**
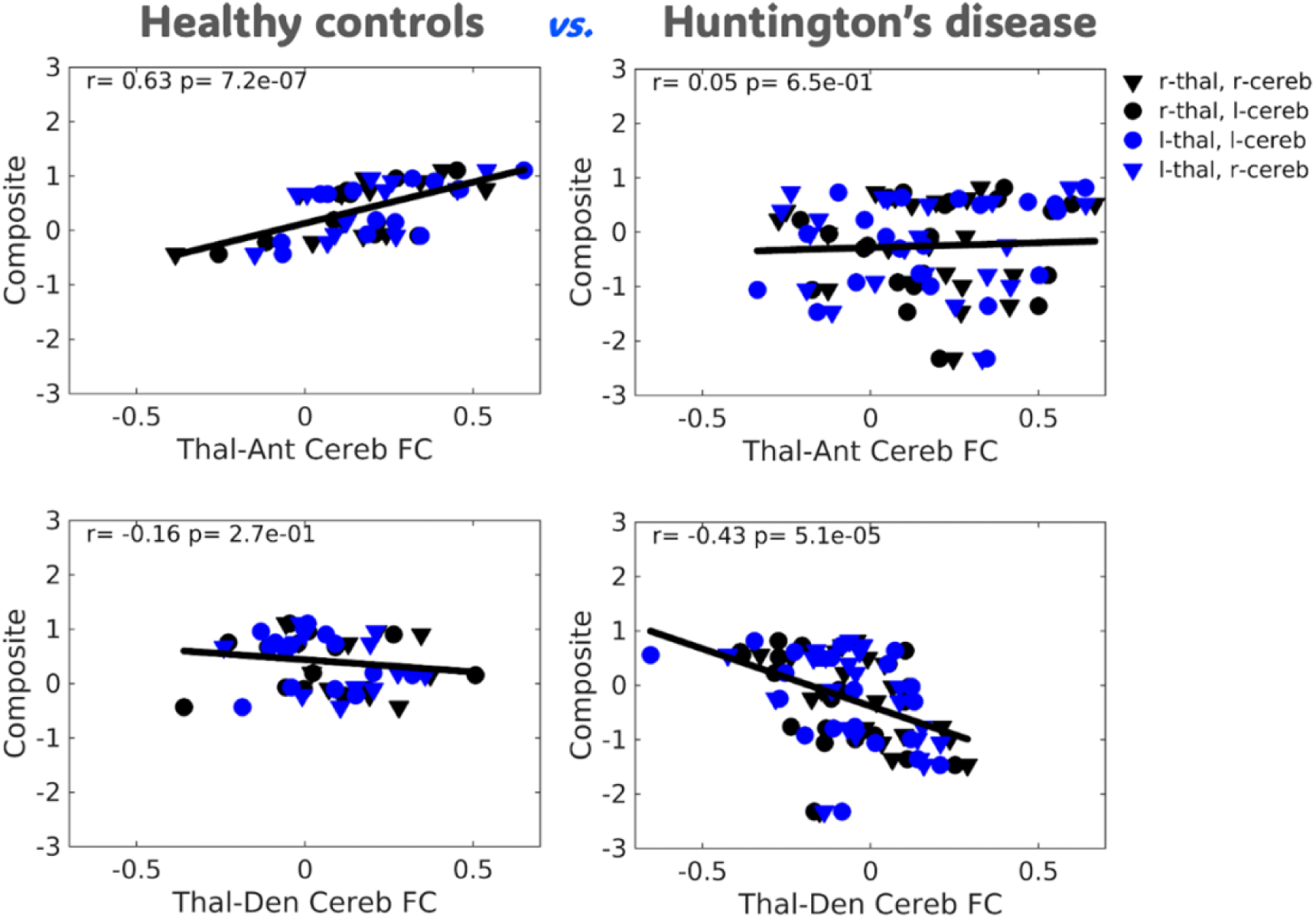
Relationship between cognitive performance and thalamo-cerebellar connectivity in 7T HD patients versus controls. Replicating that shown in Figure 3B of the main text, when patients and controls are evaluated separately, cognitive performance (a composite measure of performance on inhibition, task switching, executive function, and visuospatial judgment tasks) remains positively correlated to thalamo-anterior cerebellar (Ant Cereb) functional connectivity (FC) and negatively correlated to thalamic thalamo-dentate nucleus (Den Cereb) FC.

**Supplementary Figure 4.**
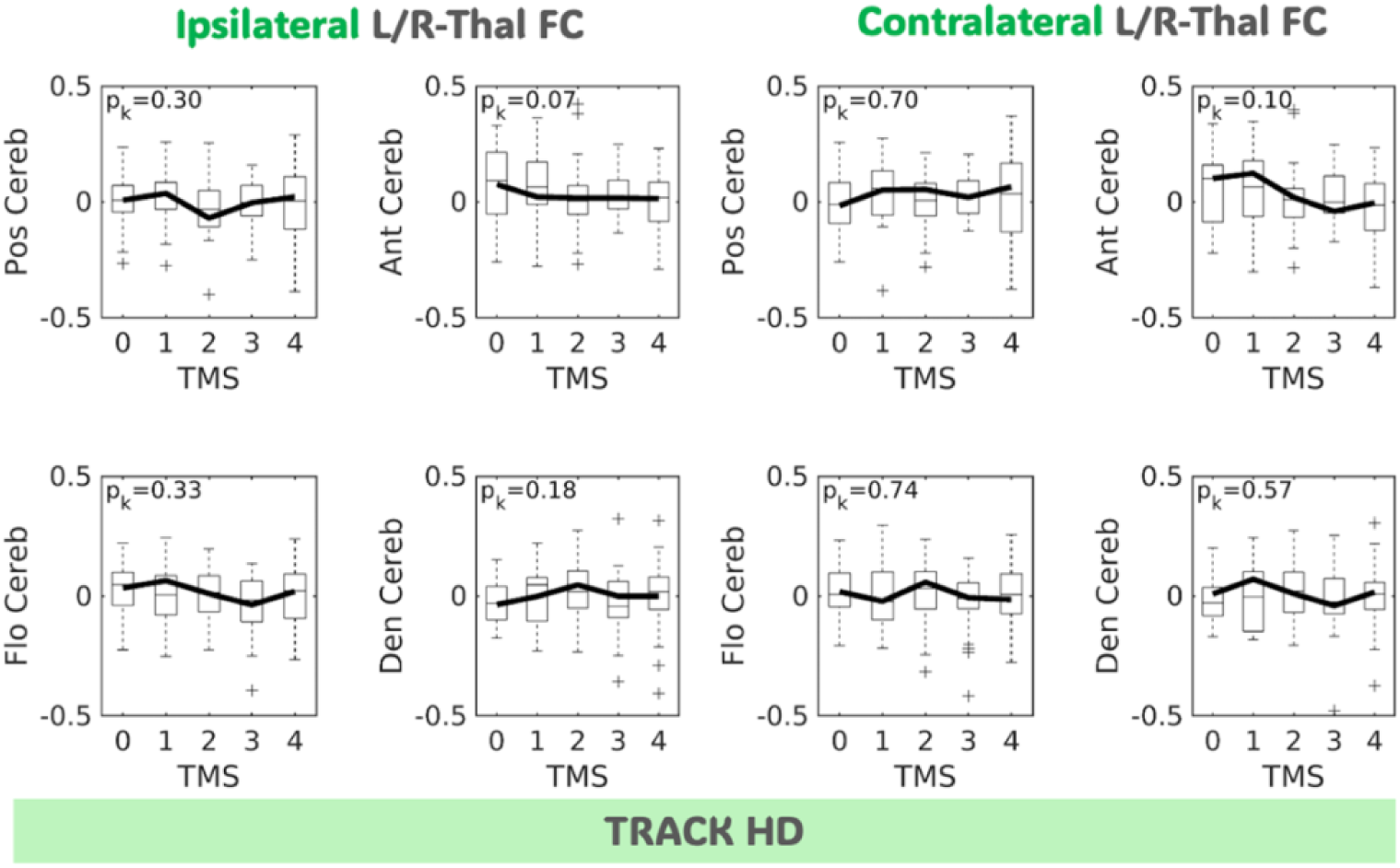
TRACK HD group differences in thalamo-cerebellar connectivity. Patients were separated into five groups based on their total motor scores (TMS) ranging 0 to 4. No significant group differences were observed, though multiple trends in the data showed transient changes in connectivity with greater disease severity or increasing TMS. HD=Huntington’s disease; l= left; r= right; Ant Cereb=anterior lobe; Pos Cereb=posterior lobe; Flo Cereb=flocculonodular lobe; Den Cereb=dentate nucleus

**Supplementary Figure 5.**
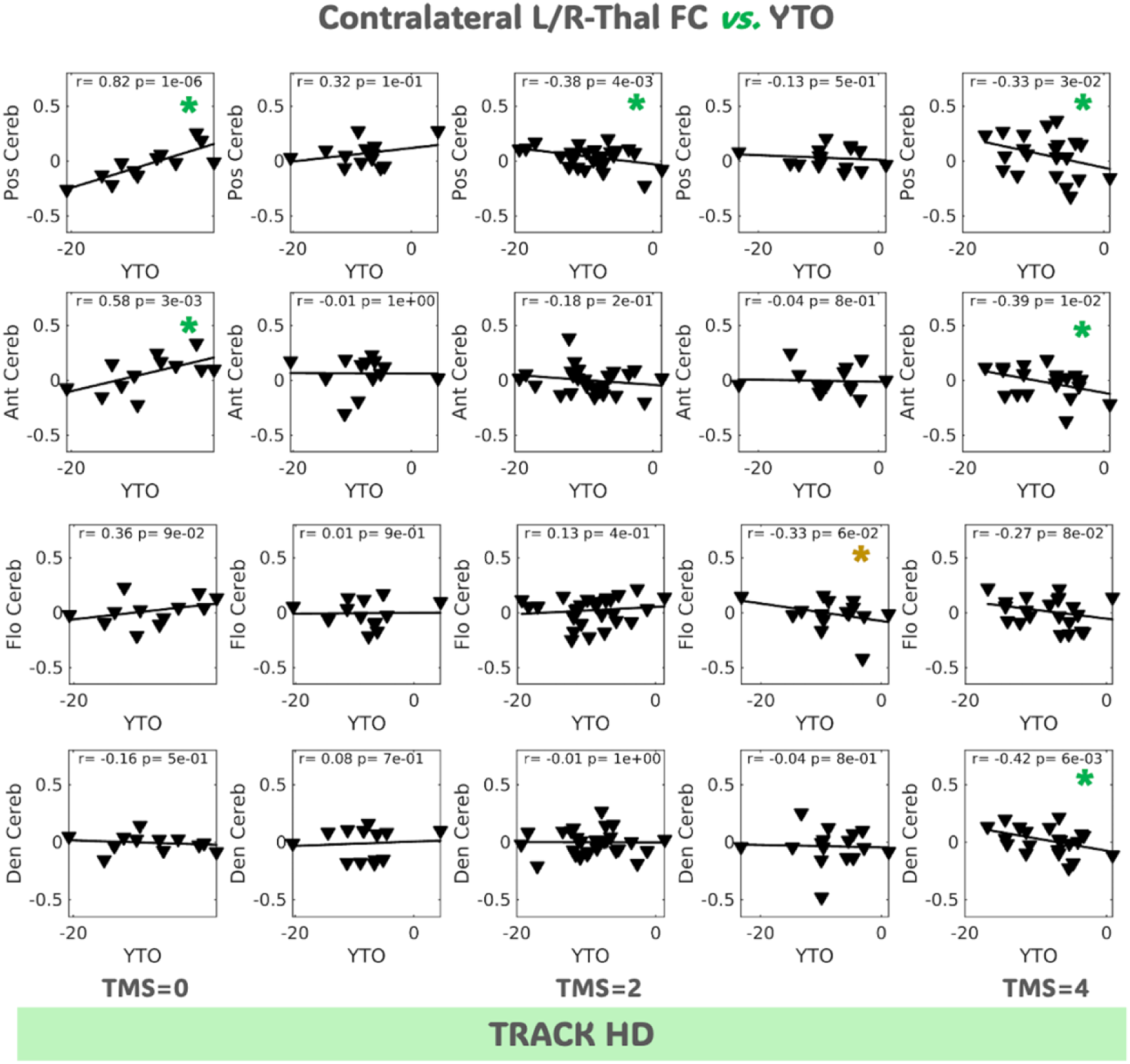
Relationship between thalamo-cerebellar connectivity and years to onset in TRACK HD cohort. Patients were separated into five groups based on their total motor scores (TMS) ranging 0 to 4. As observed for ipsilateral connections in Figure 4 of the main text, distinct correlative trends, some significant (as denoted by the green stars) or near-significant (gold star), were observed for patients with low versus higher TMS. FC=functional connectivity; Thal=thalamus; HD=Huntington’s disease l= left; r= right; Pos Cereb=posterior lobe; Ant Cereb=anterior lobe; Flo Cereb=flocculonodular lobe; Den Cereb=dentate nucleus

